# Modestly protective cytomegalovirus vaccination of young children effectively prevents congenital infection at the population level

**DOI:** 10.1101/2022.03.24.22272898

**Authors:** Catherine Byrne, Daniel Coombs, Soren Gantt

## Abstract

A vaccine to prevent congenital cytomegalovirus infection (cCMV) is a public health priority. cCMV results from maternal primary or non-primary CMV infection (reinfection or reactivation of chronic infection) during pregnancy. Young children are a major source of transmission to pregnant women because they shed CMV at high viral loads for prolonged periods. CMV vaccines evaluated in clinical trials so far have demonstrated only approximately 50% efficacy against maternal primary infection. None of these have been approved, as higher levels of vaccine-induced immunity are assumed to be required to substantially reduce cCMV prevalence. Here, we designed a mathematical model to capture the relationship between viral shedding by young children and maternal CMV infections during pregnancy. Using this model, we were able to quantify the efficacy of CMV immunity following infection to protect against reinfection and viral shedding. There was a 35% reduction in the risk of infection to a seropositive person (reinfection) versus a seronegative person (primary infection), given the same exposure. Viral shedding following reinfection was only 25% the quantity of that following primary infection. We also found that a vaccine that confers the equivalent of infection-induced immunity, when given to young children, markedly reduces both CMV transmission to pregnant women and rates of cCMV. Thus, we predict that vaccine candidates that have already been shown to be only modestly protective may in fact be highly effective at preventing cCMV by interrupting child-to-mother transmission.

## 1. Introduction

Cytomegalovirus (CMV) infects the majority of the world’ s population, and is the most common congenital infection. Congenital CMV infection (cCMV) is a major cause of neurodevelopmental abnormalities, including sensorineural hearing loss, intellectual disability, and visual impairment (1–3). Thus, creating a vaccine to prevent CMV infection in women of childbearing age has long been a top public health priority (4). However, despite promising candidates that have been evaluated or are in the development pipeline (5), no approved vaccine is yet available. CMV is a challenging vaccine target in part because prior-infection-induced immunity is insufficient to prevent reinfection or cCMV. Thus, vaccine-induced sterilizing immunity is considered to be a high bar (1).

CMV transmission occurs through exposure to breastmilk, saliva, urine or other bodily fluids of an infected individual (6). Like other herpesgroup viruses, after primary infection, CMV establishes latency and lifelong infection. Although viral reactivation occurs periodically among chronically-infected individuals, the frequency of detectable virus, and the virus load on detection, is highest following primary infection. The amount of virus shed is also age-dependent, with young children shedding CMV at much higher levels in saliva and urine and for far longer than adults (7–9). Because primary infection often occurs early in life (9–11), exposure to young children is the dominant risk factor for CMV acquisition (7,9,12–14).

cCMV can result from either a primary or non-primary maternal CMV infection during pregnancy (15,16). Transmission to the fetus occurs at the highest rates (30-60%) when primary maternal infection occurs during pregnancy (15,17). Nevertheless, most cCMV cases occur in mothers with non-primary infection, either from reinfection or reactivation of latent virus (16,18,19). CMV reinfection with another viral strain is common, including during pregnancy when it is strongly associated with the risk of cCMV (9,20–23). The relative contribution of maternal reinfection versus reactivation to cCMV transmission remains unknown, however, because of the challenges of detecting CMV reinfection (1,22). The importance of maternal reinfection is likely underestimated, as the serologic assays that have been used to estimate the frequency of reinfection only include a handful of different strains. Furthermore, although maternal immunity following infection appears to provide some protection against cCMV (24), this has not yet been well quantified.

Horizontal CMV transmission is relatively inefficient and typically appears to require repeated exposures to high viral loads (25,26). Even modest increases in immunity, or small reductions in the viral load of exposures, as would be conferred by a CMV vaccine, may therefore have a large impact on the likelihood of an individual becoming infected. Furthermore, women of childbearing age may not be the optimal target for CMV vaccination. Because infection frequently occurs early in childhood, as well as considering the fact that children appear to be the dominant source of CMV transmission (7–9,12), vaccination of younger age groups may be more effective at reducing the overall CMV prevalence and as a result, lead to fewer cases of cCMV.

To determine the optimal CMV vaccination strategy, mathematical models have been developed to describe viral transmission and simulate the use of vaccines with various characteristics, and in different target populations (14,19,27–29). While some of these models were age-structured (14,19,29), incorporated transmission of CMV from mother to infant due to breastfeeding (14,29), or accounted for potential maternal reinfection (14,19), none included the disproportionately high risk of transmission from young children to their mother, or fit parameters describing the waning of infectiousness or of post-infection immunity over time. Furthermore, none examined a vaccine that conferred an equivalent level of immune protection to that achieved following natural infection, a goal that appears achievable.

Here, we present a mathematical model of CMV transmission specifically focused on describing the age-specific infectiousness of individuals, the probability of reinfection, and the close relationship between mothers and their infants, in a resource-rich population. We used this model to quantify the immune protection from natural CMV infection, and to estimate the impact of a vaccine that induces the equivalent of post-infection protection on the burden of cCMV in the population. Finally, we compare such a vaccine with those CMV vaccines (30–32) that have already been evaluated in clinical trials but have not been approved for use.

## 2. Methods

### 2.1 Model design

We designed a stochastic microsimulation model describing the infection dynamics of CMV in a population of approximately 10,000 individuals. The estimated rates of interactions among individuals are both age and sex-specific, and are derived from the POLYMOD study contact matrix (33,34). In addition to these standard interactions, we also accounted for behaviours of mothers and their children as well as transmission of CMV through ingestion of breast milk or contact with urine in diapers (Figure 1 A).

**Figure 1:**
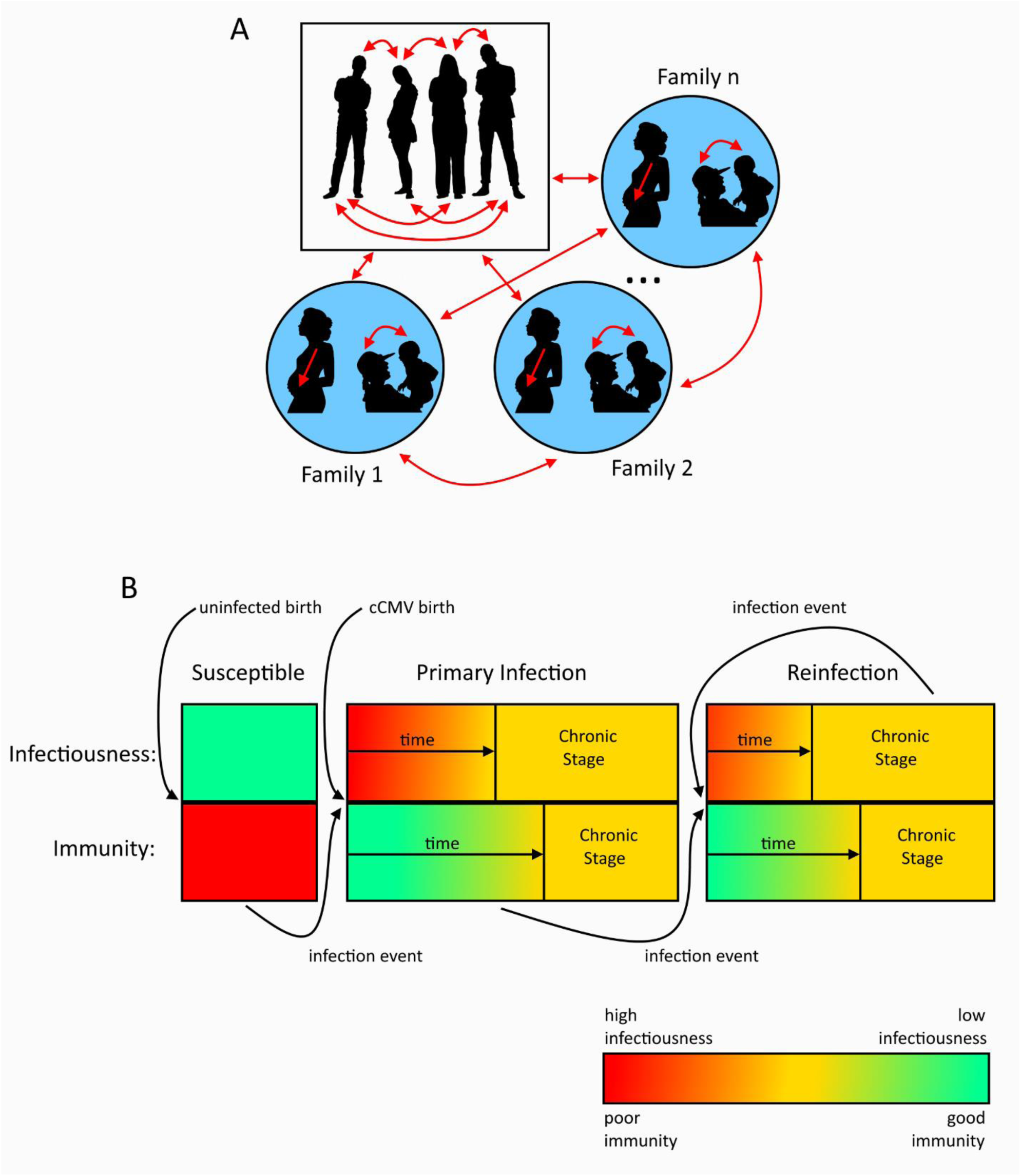
Description of CMV transmission within a population and an individual’ s infection-state transition. **Panel A** shows the different routes of transmission within a population. Transmission can occur from mother to fetus during pregnancy, due to close contacts and behaviours that result in exposure to bodily fluids among the members of the population, or specifically between mother and infant during breastfeeding or diaper changing. **Panel B** shows how individuals can progress through the stages of being susceptible, having primary infection, or having reinfection. The levels of infectiousness and immunity are quantified throughout each stage. Infectiousness and immunity are boosted following each infection event and wane over time.

Figure 1 B shows an individual’ s possible transitions between infection states. CMV-naïve individuals are categorized as being susceptible to primary infection. Individuals can then move from being susceptible to having a primary infection if CMV transmission occurs, and then, over time, to a chronic stage. Infants born into the model are categorized as either susceptible, or in the primary infection stage if they have cCMV. Individuals in any stage can move into the reinfection stage, and reinfection can occur repeatedly. During each stage of infection, the model tracks both the infectiousness, defined as the per-contact rate at which someone infects a susceptible individual, and the level of immune protection individuals have against CMV over time. Thus, an individual’ s probability of (re)infection is determined by three factors: their level of immunity, their rate of contact with other individuals, and the average infectiousness of their contacts.

CMV-uninfected individuals are not infectious and have a low immune barrier to acquiring infection. If infection occurs, CMV-uninfected individuals move into the primary infection class. During primary infection, individuals are initially highly infectious and have a high level of immunity against incident reinfection; however, both values wane over time until they reach intermediate, chronic levels. At any time, these individuals may become reinfected during interactions with other infectious individuals and enter the reinfection class. Reinfection behaves similarly to primary infection with respect to increased immunity and infectiousness that each wane over time. Because pre-existing immunity is associated with reduced viral shedding, however, we assume infectiousness during reinfection is lower than during primary infection. We also assume that since young children are known to shed more virus than adults, infectiousness depends on the age at the time infection is acquired.

We implemented the model stochastically using the tau-leaping algorithm, progressing through time steps of 1 month (35). At every time step, we calculated the probability that each individual becomes infected and randomly selected infection events to occur from an Euler-multinomial distribution. Further details on model structure, with equations and information on the implementation, can be found in the Supplementary Materials.

### 2.2 Model fitting

We fit unknown parameters of our model using Approximate Bayesian Computation (ABC), following the Lenormand Algorithm (36) to data on cCMV prevalence and from the NHANES study describing CMV seroprevalence across the ages 1-49 years old (36,37). Further details can be found in the Supplementary Materials.

### 2.3 Simulating vaccination

We simulated delivering either a vaccine that induces the equivalent of immunity due to natural infection, or an idealized vaccine that provides complete, life-long sterilizing immunity against CMV infection/reinfection. We simulated delivering these vaccines to either individuals who were 2 months, or 2, 12 or 25 years old, and assumed either 33%, 67% or 100% vaccine uptake.

We also analyzed the impact of booster doses of the vaccine that confers the equivalent of infection-induced immunity. We examined scenarios where either one or two boosters were given at intervals matching the half-life of post-infection immunity estimated by our model. We also examined scenarios where boosters were given once the child reached either 2 years old or 12 years old. For all vaccine scenarios, including boosters, we assumed that all individuals who received the initial vaccine adhered to receipt of the booster(s).

Finally, we simulated a clinical trial with the vaccine confering the equivalent of infection-induced immunity. Here, we mimicked the conditions of randomized CMV vaccine efficacy trials for which results are available (30,31). Either placebo or a single dose of the vaccine was administered to CMV-uninfected individuals, with 250 people in each group, and we observed CMV-seroconversion over a 42-month period. We performed simulations where either non-breastfeeding new mothers with infants under the age of 1 year old, women of child-bearing age, or 2-month-old children were randomized in the vaccine trial. Vaccine efficacy was defined as

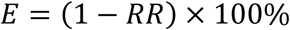

where *E* is the efficacy of the vaccine and *RR* is the risk ratio when comparing the number of individuals who acquired CMV in the placebo and vaccine groups.

## 3. Results

### 3.1 Mathematical model fits data on CMV prevalence with high accuracy

Results from model fitting are shown in Figure 2. The best-fitting simulations predicted a median cCMV rate of 0.78% (IQR 0.64-0.99%) live births (Figure 2 A), with a median percentage error of 31% (IQR 12-65%), which is similar to previous estimate (38,39). Simulations also reproduced the prevalence of CMV across ages well, as compared to the United States NHANES study, with a median percentage error of 4.0% (IQR:3.5-4.6%) (Figure 2 B).

**Figure 2:**
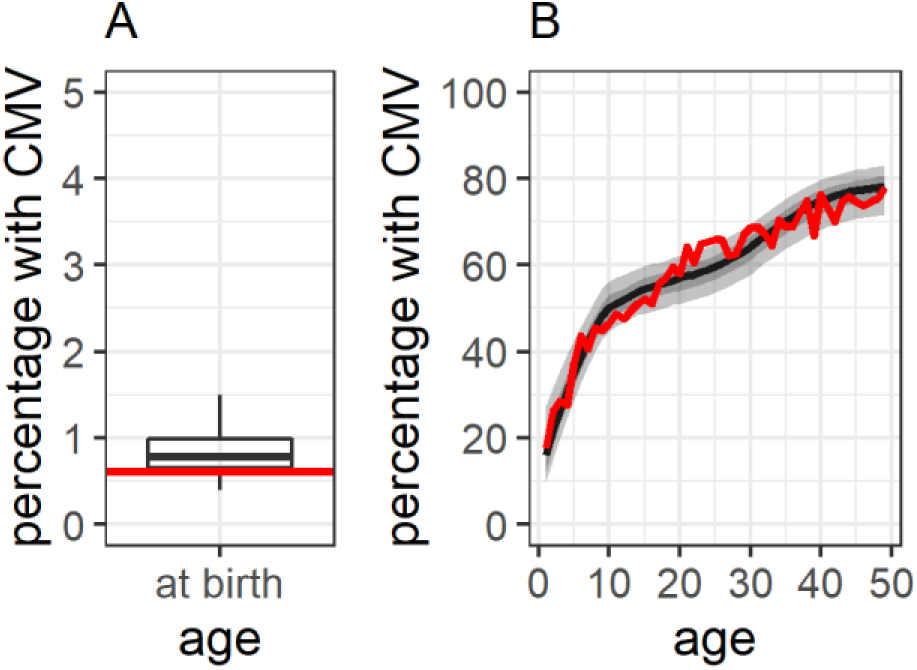
Model simulations match data on CMV prevalence well after fitting. Model-predicted CMV prevalence at birth is shown in **panel A**, and across all ages in **panel B**. In both graphs, black lines show the median value. 0.05, 0.25, 0.75 and 0.95 quantiles are also shown with a boxplot in **A** and ribbons in **B**. US NHANES CMV prevalence data across ages 1-49 years **(37)** is shown in the red line.

### 3.2 Model-predicted parameter values describing the level of immunity and infectiousness possessed by infected individuals

Figure 3 shows the distributions of the model parameters that were fit to the data using ABC. The per-contact infectiousness of adults and children immediately following primary infection, and during chronic infection are shown in Figure 3 A. Figure 3 B shows the factor change in infectiousness during reinfection as compared to primary infection. These parameter distributions show that, immediately following infection, children are on average 11.32 (IQR: 5.77-14.97) times more infectious than acutely infected adults. Similarly, individuals are on average 75% (IQR: 63-84%) less infectious during a reinfection than a primary infection. Chronically-infected individuals are substantially less infectious, being on average 2,641 (IQR: 609-3,370) times less infectious than children immediately following primary infection, or equivalently, 254 (IQR: 66-318) times less infectious than adults immediately following primary infection.

**Figure 3:**
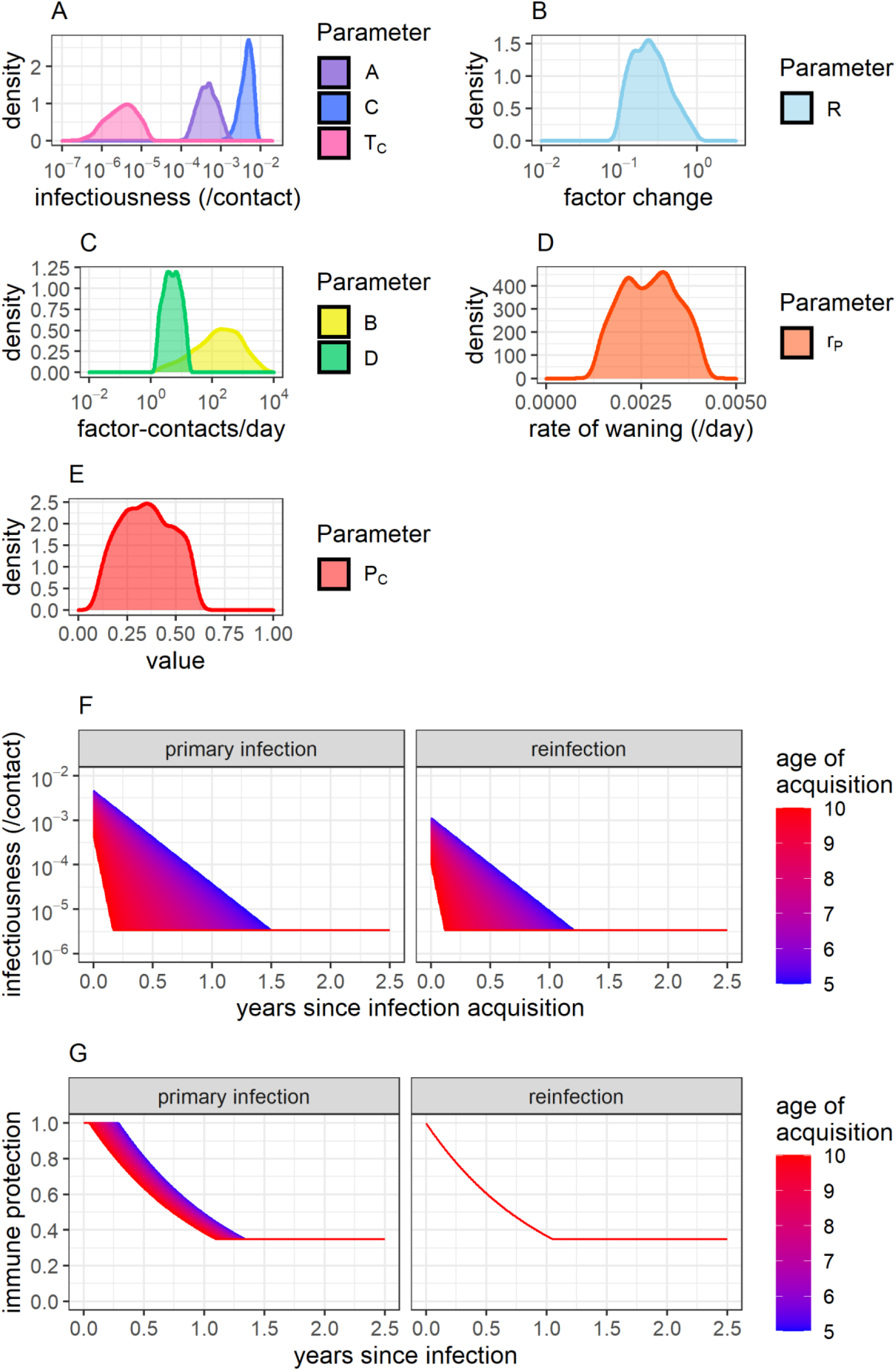
Model parameter distributions and description of immunity and infectiousness. Distributions for parameters capturing the infectiousness of adults and children immediately following primary infection (A and C) and the infectiousness of individuals in the chronic stage of infection (***T***_***C***_) are shown in **panel A**. The factor by which infectiousness is decreased during reinfection (R) is shown in **panel B**. The parameters capturing transmission of CMV between mothers and infants are shown in **panel C**. Parameters relating to the rate of immune waning (***r***_***P***_) and the level of immune protection against new infection during the chronic stage of infection (***P***_***C***_) are shown in **panels D and E**, respectively. The expected level of infectiousness (**panel F**) and immunity against reinfection (**panel G**) in individuals in the years following natural infection acquisition, stratified by the age of infection acquisition, is also shown.

With respect to transmission between mothers and their infants (Figure 3 C) our model predicts that, independent of infectiousness, there is a greater potential for infection transfer from mother to infant through breastfeeding (B) than from infant to mother through contact with diapers (D). Because the physical interactions between mothers and infants typically involve more frequent contact with infectious bodily fluids than between individuals in the general public, the per-contact probability of viral transmission is assumed to be higher. Thus, parameters B and D are reported in units of contacts/day multiplied by a factor-increase in transmission probability. On average, B is 107.63-times (IQR: 9.34-113.42) greater than D. A breastfeeding infant has an average per-day potential of CMV infection from its mother that is 305.97-times higher than through contact with an individual in the general public, while the potential for transmission from an infant to its mother due to the handing of diapers is on average 3.96-times higher per day than through contact with an individual in the general public.

In addition to reductions in infectiousness, post-infection immunity is assumed to also reduce the risk of (re)infection acquisition. The level of an individual’ s immune protection against CMV infection is represented by parameter ρ *ϵ*[0,1], where 0 implies no protection against infection and 1 implies complete protection against infection. As shown in Figure 3 D, this protection is expected to wane at a rate *r*_*p*_, averaging 0.0027/day (IQR: 0.0021-0.0033) and leading to a half-life of 275 (IQR: 212-325) days. Waning continues until reaching parameter value *P*_*C*_ (Figure 3 E), averaging 0.35 (IQR: 0.25-0.46) based on model fitting.

Using the mean values of these parameter distributions, we plotted the change in the per-contact infectiousness and level of immunity of individuals in the time following infection acquisition, as shown in Figure 3 F and G. Because young children are more infectious for a longer time than older individuals, graphs are stratified by age.

### 3.3 cCMV likelihood as determined by pregnant women’ s infection status

We determined how likely women are to experience a primary infection or reinfection during pregnancy and the probability of vertical CMV transmission for each infection stage (Figure 4 A). From our analysis, our model predicts that 64.3% of women remain in the chronic stage of infection throughout pregnancy. Our model also predicts that 3.9% of uninfected women experience a primary infection during pregnancy, which is slightly higher than the 2.3% annual seroconversion rate (range 1-7%) among pregnant women measured in cohort studies (40). In addition, 4.3% of seropositive women are reinfected during pregnancy. Importantly, of all incident infections (primary or reinfection) that occur during pregnancy, 87% are transmitted by children, which is consistent with clinical studies (7–9,12).

**Figure 4:**
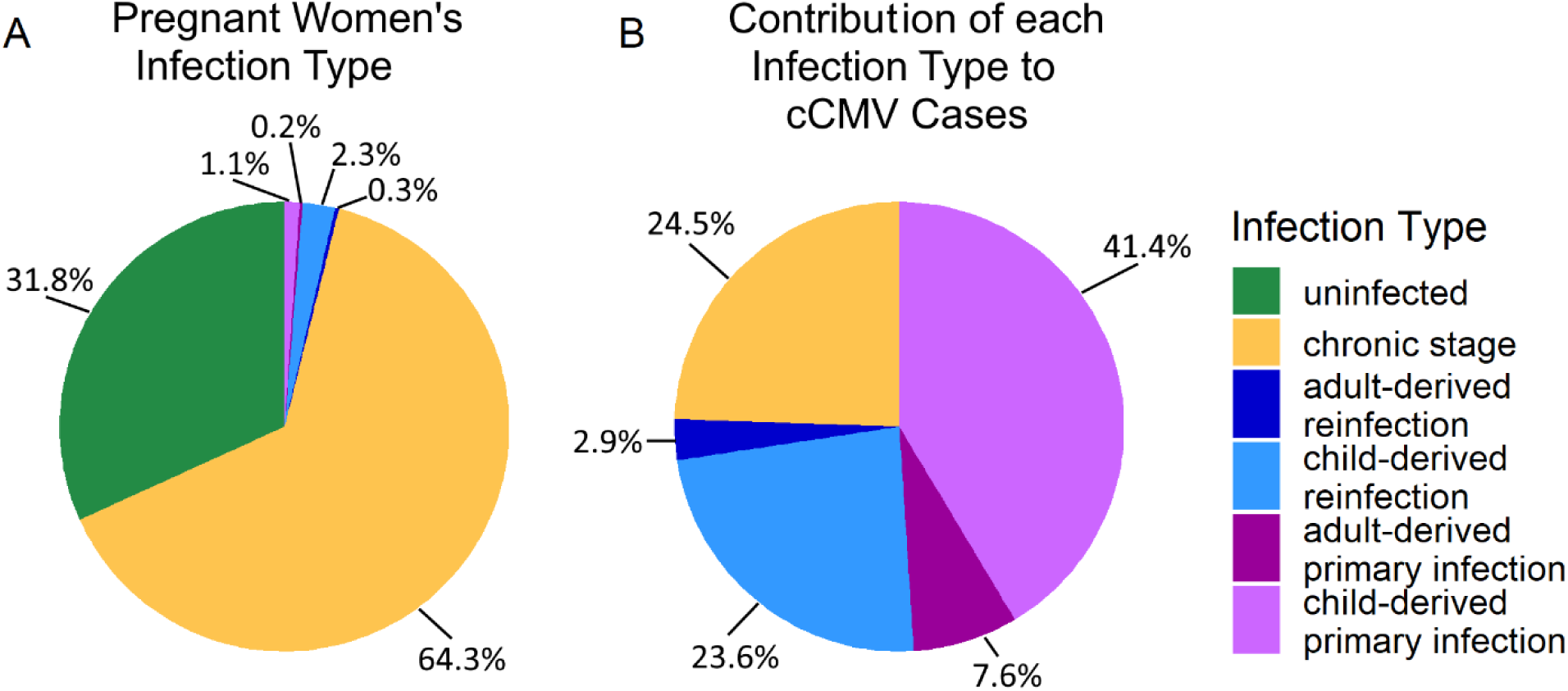
The distribution of women’ s infection statuses during pregnancy. **Panel A** shows the percentage of women in each infection stage during pregnancy, and **panel B** shows the percentage of cCMV cases attributable to each infection type. Individuals in the chronic stage may have been infected once, or multiple times, in the past. Active infections are stratified based on the source of infection (acquired either from children or adults).

In our model, we assumed that the risk of CMV transmission to the fetus during pregnancy was dependent on the type of infection and level of immunity (primary, reinfection, or chronic). We fixed a 33.4% risk of vertical CMV transmission due to primary maternal infection during pregnancy throughout model simulations, based on clinical data (19). In comparison, our model predicted a median risk of vertical CMV transmission of 8.2% (IQR:5.5-12.2%) among women reinfected during pregnancy, and 0.2% (IQR:0.10-0.51%) due to reactivation of chronic maternal infection. Overall, our model predicted that 51.0% of cCMV were due to non-primary maternal infection; of these cases, 52.0% were related to reinfection during pregnancy and 48.0% to chronic infection (Figure 4 B). The remaining 49.0% of cCMV cases resulted from primary maternal infection, higher than the 25-34% estimated by previous models (16,18,19).

### 3.4 Vaccination that induces sterilizing immunity

We first examined the effects of an idealized vaccine that confers life-long sterilizing immunity against CMV. While such a vaccine is improbable, this analysis offers a best-case scenario with which to compare other, more realistic vaccines. Figure 5 A shows the decline in cCMV cases in the 50 years following vaccine implementation while Figure 5 B shows how much each infection status in pregnant women contributes to overall cCMV case numbers.

**Figure 5:**
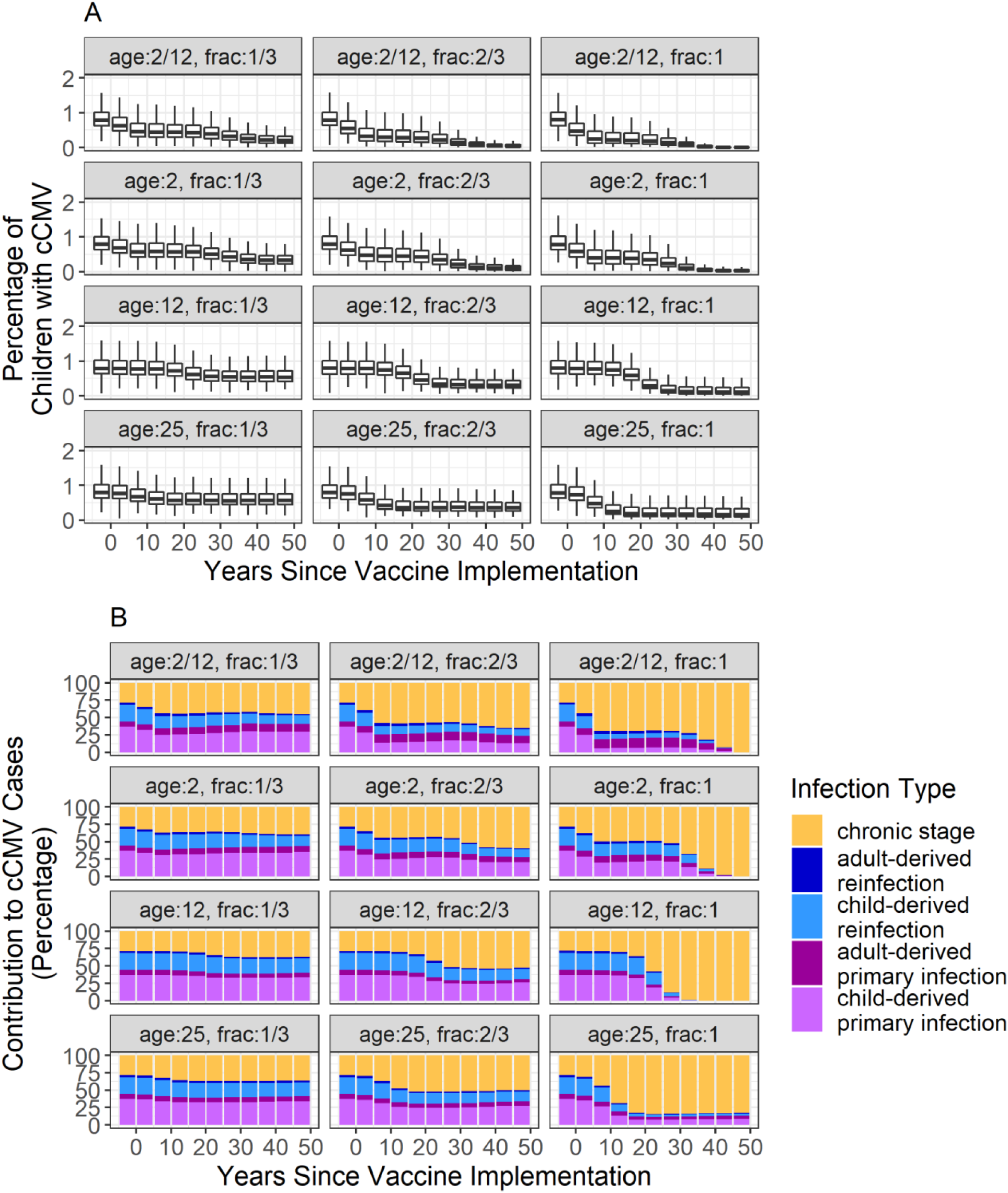
cCMV cases following implementation of a vaccine that confers sterilizing immunity and the contribution of each infection type. Data are stratified by the fraction of the population vaccinated and the age in years at which vaccines are given. **Panel A** shows the percentages of children born with cCMV following vaccine implementation. The dark middle line of each boxplot, the height of the boxes, and whisker lines represent the median, interquartile range, and range, respectively. In **panel B**, the mean percentage of cCMV cases due to different infection statuses in pregnant women is depicted by the size of the coloured bars. Note that vaccinated women acquiring their first infection during pregnancy are classified here as having a reinfection.

Young ages at vaccination, such as at 2 months and 2 years old, led to marked declines in the percentage of cCMV cases. When the vaccine is administered to 2-month-old children, our model predicts cCMV cases are expected to drop by 74.6%, 95.4%, or 99.4% over the course of 50 years when vaccination coverage is 33%, 67%, or 100%, respectively (Figure 5 A). As the target age increases, the benefit of vaccination decreases. When vaccinating 25-year-olds, the oldest age group examined, there is little change in the percentage of children born with cCMV; dropping by only 27.4%, 54.5%, or 80.0% over the course of 50 years when vaccinating 33%, 67%, or 100% of 25-year-olds, respectively (Figure 6 A).

**Figure 6:**
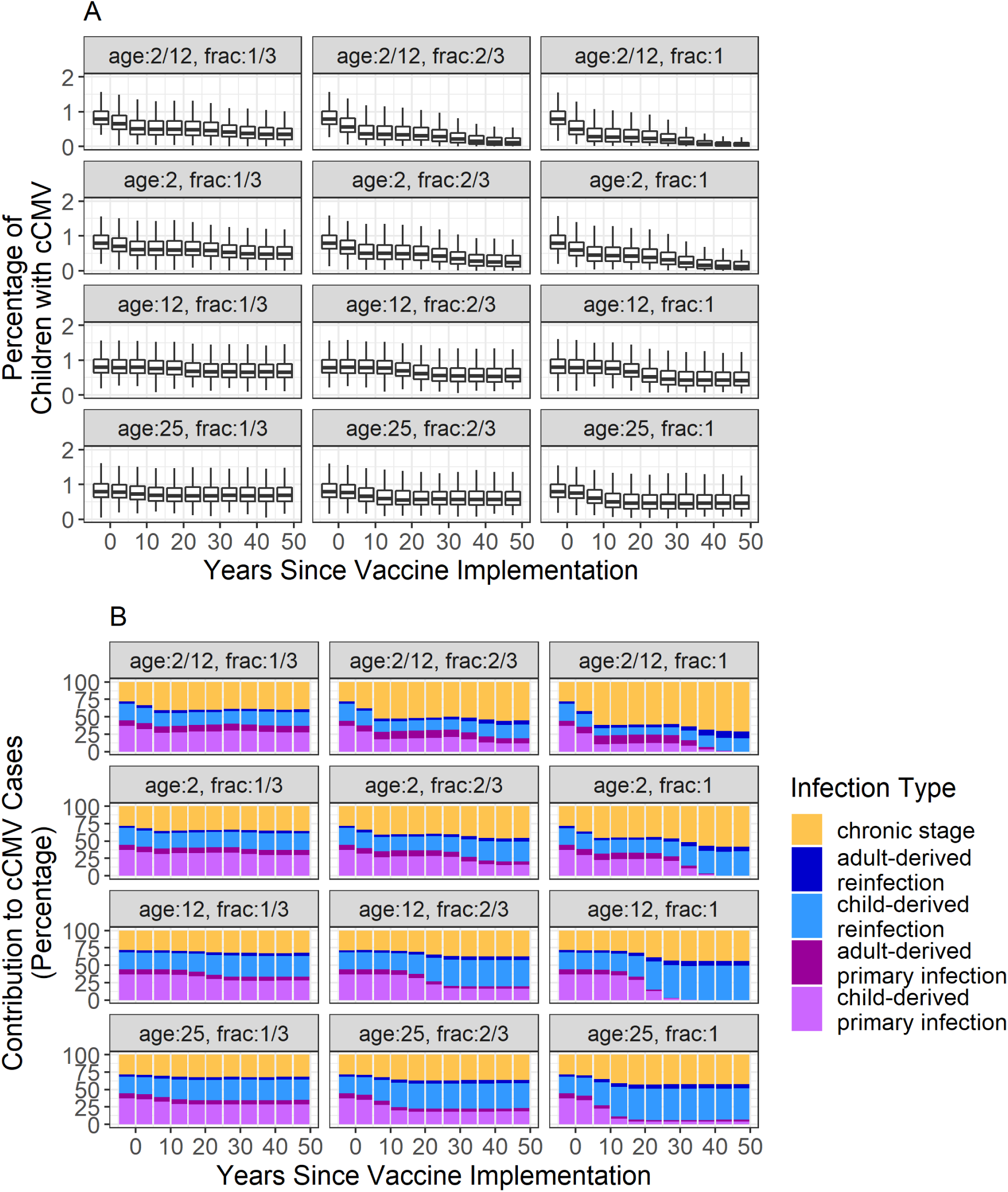
cCMV cases following implementation of a vaccine that confers the equivalent of post-infection immunity and the contribution of each infection type. Data are stratified by the fraction of the population vaccinated and the age in years at which vaccines are given. **Panel A** shows the percentages of children born with cCMV following vaccine implementation. The dark middle line of each boxplot, the height of the boxes, and whisker lines represent the median, interquartile range, and range, respectively. In **panel B**, the mean percentage of cCMV cases due to different infection statuses in pregnant women is depicted by the size of the coloured bars. Note that vaccinated women acquiring their first infection during pregnancy are classified here as having a reinfection.

These declines can be directly linked to the changes in the infection statuses of pregnant women (Figure 5 B and Figure S. 2 of the Supplementary Materials). Early childhood vaccination markedly decreases CMV transmission from young children to women (Figure S. 2 of the Supplementary Materials), leading in turn to overall reductions in the number of cCMV cases. Within 50 years post-vaccine implementation, primary CMV infections in pregnant women, dropped from causing 49.0% of cCMV cases to causing an average of 41.0%, 23.4% or 0.0% of cCMV cases when vaccinating 33%, 67% or 100% of 2-month-olds, respectively (Figure 5 B). Alternatively, when vaccinating 25-year-olds, many pregnant women still acquired new infections during pregnancy (Figure S. 2 of Supplementary Materials). As such, primary CMV infection in mothers still accounted for an average of 41.0%, 33.7% and 13.3% of cCMV cases when vaccinating 33%, 67% or 100% of individuals, respectively (Figure 5 B).

In addition to preventing cCMV, a vaccine is also expected to reduce the number of infected individuals within a population overall. Thus, we calculated the effective reproductive value (R_eff_), defined as the number of other infections caused by one infected individual over the course of their lifetime. A full description of the calculation of R_eff_ is given in the Supplementary Materials, with resulting changes due to vaccination shown in Figure S. 3. As the number of CMV cases within resource-rich populations is assumed to be constant over time, R_eff_ equals 1, on average, in the absence of vaccination. Vaccinating 25-year-olds only decreased R_eff_ values by either 1.7%, 3.4%, or 4.6%, 50 years post-vaccine implementation, with 33%, 67%, or 100% uptake in the population, respectively. However, vaccinating 2-month-olds decreased R_eff_ values by 22%, 54%, or 75% 50 years post-vaccine implementation, with 33%, 67%, or 100% of uptake, respectively.

### 3.5 The impact of vaccination that confers immunity comparable to natural infection

Although the development of a vaccine that confers potent and long-lasting sterilizing immunity to CMV has been challenging, modest vaccine-mediated protection against primary infection has already been demonstrated (30,31). Furthermore, evidence indicates that CMV vaccines can readily induce immune responses comparable to seropositive individuals (30,31,41). As such, we examined the effects of a vaccine that mimics the immunity induced by natural infection, i.e., with the characteristics shown in Figure 3 F and G. Results from a vaccination that provides immunity equivalent to that achieved post-natural infection are shown in Figure 6.

As with the idealized vaccine, the greatest reduction in cCMV from a vaccine that mimicks post-infection immunity is obtained by targeting young infants, and this benefit decreases with age at vaccination (Figure 6 A). This is also true with respect to the changes in pregnant women’ s infection statuses (Figure 6 B and Figure S. 4 of the Supplementary Materials) as well as declines in R_eff_ (Figure S. 5 of the Supplementary Materials). Since the immunity induced by this imperfect vaccine (akin to that by natural infection) wanes significantly over time (Figure 3 G), we also examined the effects of booster doses. In none of the booster scenarios was there a noticeable decline in the rate of cCMV compared to the base case of a single vaccination. Results of simulations with boosters can be found in Figure S. 7 of the Supplementary Materials. A direct comparison of the outcomes for a vaccine providing sterilizing immunity and a single dose of a vaccine providing the equivalent of post-infection immunity, each given at 2 months of age, can be found in Table 1.

**Table 1:** Comparison of CMV outcomes 50 years post-implementation of vaccination. Reduction in cCMV cases and **R**_**eff**_ values are shown with use of a vaccine that provides life-long sterilizing immunity, or one that confers the equivalent of post-infection immunity, at 2 months of age and assuming different levels of vaccine uptake.

| Measure, 50 years post vaccine implementation | % target population vaccinated | Immunity induced by vaccine |  |
| --- | --- | --- | --- |
|  |  | Life-long sterilizing immunity | Post-infection immunity |
| % reduction in cCMV prevalence | 100 | 99.4% | 94.9% |
|  | 67 | 95.4% | 86.2% |
|  | 33 | 74.6% | 56.2% |
| $R_{eff}$ | 100 | 0.016 | 0.278 |
|  | 67 | 0.205 | 0.501 |
|  | 33 | 0.626 | 0.846 |

To better understand the impact of vaccinating young children on cCMV cases, we examined the time of infection and infectiousness of children under the age of 5, who are most likely to infect pregnant women due to a combination of their high viral shedding in saliva and urine as well as their types of interactions. In the absence of vaccination, our model predicts CMV infection in this group occurs at a mean age of 1.86 years (IQR: 1.59-2.17). This explains why vaccinating at 2 months of age provides greater protection against CMV than vaccinating at 2 years, by which time most pre-school age infections have already occurred. Following administration of a vaccine that provides the equivalent of post-infection immunity, our model predicts that vaccinated children will still have breakthrough infections. However, the quantity and duration of viral shedding of these children with breakthrough infection is reduced compared to infected, unvaccinated children. As such, on average, vaccinated children are markedly less infectious to pregnant women. Further information showing the impacts of vaccination on childhood infection status and infectiousness can be found in Figure S. 6 of the Supplementary Materials.

### 3.6 Comparing results to existing vaccines

To simulate a clinical trial testing a vaccine that confers the equivalent of post-infection immunity, and compare these to completed CMV vaccine trials, we reproduced the design and conditions of the original phase 2 glycoprotein B/MF59-adjuvanted vaccine trial (30), and the Merck V160 vaccine trial (31). As in those trials, we randomized either CMV-seronegative women within 1 year of giving birth (30), or seronegative women of child-bearing age (31), to receive either a placebo or a vaccine inducing the equivalent of post-infection immunity. Given our previous finding, we also simulated a trial in which seronegative, male and female 2-month-olds were randomized. Results are shown in Figure 7.

**Figure 7:**
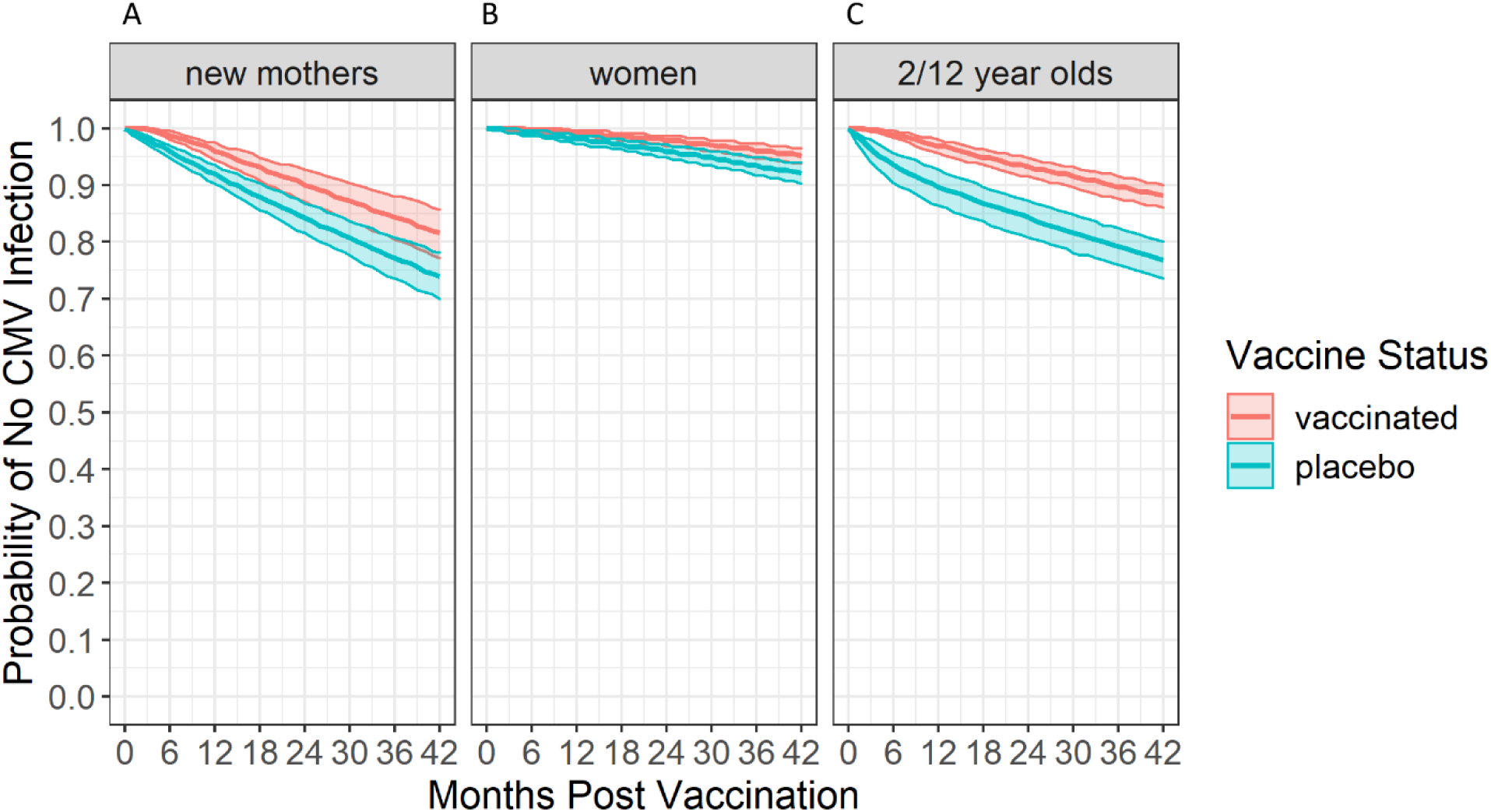
Simulated clinical trial results for a vaccine that induces the equivalent to post-infection immunity against CMV. The probability of no CMV infection in the months following the administration of either the vaccine or a placebo to different target groups are shown. Panel A shows results when seronegative women within 1 year of giving birth were randomized, as in the design of Pass, et al. **(30). Panel B** shows results when seronegative women of child-bearing age were randomized, as in the design of Das, et al. **(31). Panel C** shows results when seronegative 2-month-old children were randomized. Coloured lines show the median probability of no CMV infection across simulations while ribbons show the interquartile range.

Over 42 months of simulated study follow-up, vaccine efficacy against incident CMV infection was 28.9% (IQR: 18.5-40.0%) when given to new mothers, less than the 50% observed with the glycoprotein B/MF59-adjuvanted vaccine (30). Similarly, vaccine efficacy was 39.4% (IQR: 20.0-34.0%) in all women of child-bearing age, compared to 42.4% with the V160 vaccine (31). Finally, an efficacy of 46.9% (IQR: 38.2-56.7%) for a vaccine inducing the equivalent of post-infection immunity was observed among 2-month-old children.

## 4. Discussion

A better understanding of immune protection against CMV infection and transmission is essential for informing the development of a vaccine to reduce the enormous burden of cCMV (1,5,14). The rate of primary maternal CMV infection in pregnant women and their risk of transmitting to the fetus is well described (1,5). However, because CMV reinfection cannot be reliably diagnosed (1,22), the frequency of maternal reinfection during pregnancy, the effectiveness of post-infection immunity against incident (re)infection, and the relative contribution of reinfection (compared to reactivation or chronic infection) to cCMV are central questions in the field. As such, we designed a comprehensive stochastic microsimulation model of CMV transmission in a resource-rich population, which allowed the estimation of the rate of CMV reinfection during pregnancy and, as a corollary, the risk of congenital transmission associated with maternal reinfection.

Using this model, we predicted that, on average, 4.3% of seropositive women are reinfected during pregnancy, which is lower than the rate of approximately 30% previously estimated in high prevalence populations (22), but higher than that of primary maternal infection (40). For the first time, we also quantified the level of immunity following infection with respect to the relative risk of primary infection versus reinfection, which has a half-life of around 9 months, but still decreases long-term acquisition risk by at least 35% compared to naïve individuals. Furthermore, we estimated that pregnant women with a CMV reinfection transmit to their fetus 8.2% of the time, compared to >30% with primary maternal infection. Interestingly, although the risk of cCMV was only 0.2% among women with infection that remained chronic during pregnancy, such women accounted for nearly one-quarter of all cCMV cases. Altogether, these results highlight the importance of preventing all maternal CMV infections through vaccination (1,24,42).

Developing an effective CMV vaccine has long been a top public health priority, but achieving a vaccine that induces a high level of sterilizing immunity has proven to be a difficult goal to achieve (5). However, despite its ubiquity, CMV infection appears to be rather inefficient; for example, on average, transmission to infants through breastfeeding requires months of exposure to virus in breast milk (25,26,43,44). Thus, we hypothesized that a vaccine that provides even modest improvements to immunity could still prove beneficial. Furthermore, given that our model indicates that 87% of primary infections and reinfections during pregnancy are from viral shedding by young children, we hypothesized that even a modest reduction of infectiousness early in life would have outsized benefits for preventing cCMV (17,45,46). To test these hypotheses, we used our model to estimated the efficacy of a CMV vaccine that mimics post-infection immunity. Our results indicate that such a vaccine, i.e., one that does not show strong protection against incident infection, would nevertheless be highly effective in preventing cCMV, by up to 86.2-94.9% if there were high uptake by young children. While many breakthrough infections still occur, they are on average less infectious compared to primary infections in unvaccinated children, resulting in an outsized reduction on transmission to women. Indeed, we found that weakly protective vaccination of young children would have a greater impact on population-level cCMV rates than a targeting adolescents or young adults with a 100% protective vaccine.

Previous mathematical models of CMV vaccination have suggested that if vaccination is given too early, waning immunity may render women vulnerable during their childbearing years (28,29). However, our model indicates that the benefits of having higher immunity levels in young children far outweigh an impact of lower immunity in pregnant women. These earlier models did not account for the high risk of transmission from young children to women, thereby underestimating the impact of vaccinating children. Boosters provided negligible benefits in our model, likely because the first vaccine dose already obviates primary CMV infection, such that any breakthrough vaccination will behave like a reinfection, with much lower infectiousness. While boosters would increase protection against reinfection, our results indicate that transmission by children with primary infection is the main driver of cCMV in a population.

Our results support those of other modeling studies that young children are the optimal target of a vaccine to prevent cCMV (14,19,27–29). Routine vaccination of children against CMV would be primarily for the benefit of others, since CMV infection in healthy children is almost never clinically actionable. This approach, therefore, raises a number of important ethical, regulatory and acceptability questions (47). However, childhood vaccination to prevent congenital rubella represents a precedent for such a strategy (48). Furthermore, increasing evidence suggests that CMV infection may have deleterious long-term health effects, including increased all-cause mortality in the general population, which, if proven, could justify routine childhood CMV vaccination for its direct benefits (49).

Importantly, our results demonstrate convincingly that substantial decreases in the rates of cCMV may be possible with a vaccine that is only modestly protective against CMV acquisition. Of the vaccines that have been evaluated in efficacy trials, the recombinant gB/MF59 vaccine (30,50) and V160, a replication-deficient CMV vaccine (31,32), both with efficacies of 40-50%, appear to protect better than post-infection immunity, as determined by our simulated trials of a hypothetical vaccine that mimicks infection-induced immunity, under the same study conditions. In addition, we showed that such a vaccine has higher efficacy in 2-month-old children as compared to women of child-bearing age, the groups studied in the gB/MF59 and V160 vaccine trials (30,31), likely again reflecting the strong infectivity of young children. This suggests that either of these vaccines, if given to young children, could effectively reduce the prevalence of cCMV. Because the size of vaccine trials required to determine a statistically significant reduction in the rate of cCMV is prohibitively large, prevention of primary maternal CMV infection has been embraced as a surrogate outcome measure (47). To assess the potential impact of childhood vaccination on cCMV, therefore, endpoints of randomized controlled pediatric trials could compare the quantity and duration of CMV shedding following infection, and the frequency of transmission to close contacts, in addition to the rate of infection, among CMV vaccine and control groups (9).

In summary, the work presented here builds on previous clinical studies and mathematical models of CMV transmission and vaccination (14,19,27–29), and provides new predictions regarding the impact of infection-induced immunity, the relative contribution of maternal reinfection to cCMV, and the impact of vaccinating young children, on cCMV prevalence. A notable limitation of this work is the lack of detailed biological data available to inform some model parameters, including differences in infectiousness across age groups or during primary infection versus reinfection. In addition, while we included transmission between mothers and infants in the model, we could not account specifically for differential rates of transmission between other family members within a household, which could be important. By fitting the model to those biological data that are available, we were able to determine plausible estimates for these parameters; however, to the extent possible, clinical studies focused specifically on testing some of our results would strengthen our conclusions. In particular, our finding that use of even a modestly protective CMV vaccine, such as those that may have already been evaluated in adults (30,31), could strongly reduce rates of cCMV if administered to young children merits additional study. We propose that CMV vaccination of young children and its impact on viral shedding and onward transmission should be considered by researchers, industry partners, and regulators when determining the design of future trials and the requirements for licensure.

## Data Availability

All data produced in the present study are available upon reasonable request to the authors

