## Supplementary Materials for "Modestly protective cytomegalovirus vaccination of young children effectively prevents congenital infection at the population level"

a. Department of Microbiology and Immunology, The University of British Columbia, Vancouver, British Columbia, Canada, b. British Columbia Children's Hospital Research Institute, Vancouver, British Columbia, Canada, c. Institute of Applied Mathematics, The University of British Columbia, Vancouver, British Columbia, Canada, d. Department of Mathematics, The University of British Columbia, Vancouver, British Columbia, Canada, e. Département de Microbiologie, Infectiologie et Immunologie, Université de Montréal, Montréal, Québec, Canada.

### 1. CMV transmission model

We developed a mathematical microsimulation model to describe the transmission of CMV within a population of approximately 10,000 individuals existing within a resource-rich country. Individuals progress through the stages of being susceptible, acquiring a primary infection, acquiring a reinfection or being chronically infected.

#### 1.1 Contact matrix

CMV transmission occurs through interactions between individuals in the population. We assumed that the rate of contact between individuals in the population was both age and sex-specific and followed a contact matrix previously described based on data collected in the Netherlands as part of the POLYMOD study (35,36). Data from this contact matrix is available at https://github.com/mvboven/cmv-serology.

#### 1.2 Defining infectiousness

Within our mathematical model, individuals born without cCMV begin in the susceptible class (S) of infection and are assumed to have no immunity against CMV. These susceptible individuals will interact with infected individuals, and a transmission event may occur. Here, we define infectiousness as the per-contact rate at which an infected individual infects a susceptible individual. Following infection, a susceptible individual (S) is assumed to move into the primary infection ($I_P$) class. While in this class, the individual is assumed to have a per-contact infectiousness rate of $T_p$. $T_p$ is assumed to start high at a value T and then exponentially wane over time to a chronic rate of infectiousness, $T_C$ . However, since children are known to be much more infectious than adults and remain infectious for much longer, an individual's initial infectiousness, and how quickly their infectiousness wanes, are dependent on the age of the individual upon infection acquisition ($a_i$). Thus, we assume that the per-contact infectiousness rate of an individual immediately following infection is given by $T(a_i)$ where

$$T(a_i) = \begin{cases} C & , a_i < \alpha_C \\ \frac{A - C}{\alpha_A - \alpha_C}(a_i - \alpha_C) + C & , \alpha_C \leq a_i < \alpha_A \\ A & , a_i \geq \alpha_C. \end{cases}$$

Here, C is the initial per-contact infectiousness rate of children under the age of $\alpha_C$. A is the initial per-contact infectiousness rate of individuals over the age of $\alpha_A$. For individuals between the ages of $\alpha_C$ and $\alpha_A$, we assume an age-dependent linear decline in the initial per-contact infectiousness rate. As young children are known to shed CMV at much higher levels than older children and adults, we assume $A > C$ and set $\alpha_C = 5$ years and $\alpha_A = 10$ years (13,53,54).

We define $w(a_i)$ to be the time it takes for an individual's per-contact infectiousness rate to wane to the chronic rate ($T_C$) and specify the functional form

$$w(a_i) = \begin{cases} \omega_C & , a_i < \alpha_C \\ \frac{\omega_A - \omega_C}{\alpha_A - \alpha_C}(a_i - \alpha_C) + \omega_C & , \alpha_C \leq a_i < \alpha_A \\ \omega_A & , a_i \geq \alpha_C. \end{cases}$$

We set $\omega_C = 1.5$ years and $\omega_A$= 2 months (13,17,53,54).

Assuming an exponential decay in the per-contact infectiousness rate between the time of contracting primary CMV and the establishment of chronic infection, individual i with a primary infection has a per-contact infectiousness rate at time $\Delta t_i$ post-infection of

$$T_p(a_i, \Delta t_i) = \begin{cases} T(a_i)e^{-r_T(a_i)\Delta t_i} & , \Delta t_i < w(a_i) \\ T_C & , \Delta t_i \geq w(a_i) \end{cases}$$

where

$$r_T(a_i) = \frac{\ln T(a_i) - \ln T_c}{w(a_i)}.$$

If individuals become reinfected, we assume that their per-contact infectiousness rate is boosted but then wanes back to the chronic level of infectiousness $T_c$ in the same way it does during primary infection. However, because reinfected individuals appear to be less infectious than individuals with a primary infection, we scale their per-contact infectiousness rate by a factor R where $0 \leq R \leq 1$ such that the per-contact infectiousness rate of a reinfected individual, $T_R(a_i, \Delta t_i)$, becomes

$$T_R(a_i, \Delta t_i) = \begin{cases} RT(a_i)e^{-r_T(a_i)\Delta t_i} & , \Delta t_i < w(a_i) - \ln(1/R)/r_T(a_i) \\ T_C & , \Delta t_i \geq w(a_i) - \ln(1/R)/r_T(a_i). \end{cases}$$

Graphs showing the changes in $\mathbf{T_P(a_i, \Delta t_i)}$ and $\mathbf{T_R(a_i, \Delta t_i)}$ over time are presented in Figure 2 F of the main paper.

With these expressions defined, we define the per-contact infectiousness rate of individual i with infection-stage $s_i$ at time $\Delta t_i$ post transitioning into their current infection stage to be

$$\tau_i(a_i, \Delta t_i, s_i) = \begin{cases} 0 & , s_i = \text{susceptible} \\ T_p(a_i, \Delta t_i) & , s_i = \text{primary infection} \\ T_R(a_i, \Delta t_i) & , s_i = \text{reinfection.} \end{cases}$$

#### 1.3 CMV transmission between mothers and infants

Because of the close interactions between mothers and infants, we assume that there are higher rates of transmission between these pairs not accounted for in our contact matrix. Breastfeeding can increase the rate of infection transmission from the mother to the infant. If the infant is in diapers, a mother's contact with fecal matter and urine will increase the rate of infection transmission from infant to mother. To distinguish breastfeeding status, we assign each individual i to a state $b_i \epsilon \{0,1\}$ where 0 indicates they are not ingesting breastmilk and 1 indicates that they are. Similarly, to distinguish diaper status, we assign each individual i to a state $d_i \epsilon \{0,1\}$ where 0 indicates they are not in diapers and 1 indicates that they are.

As such, if a child is breastfeeding ($b_i = 1$), the rate infectiousness of the mother to the child becomes $B\tau_i$, where B incorporates both the number of breastfeeding events per day and increase of transmission due to breastfeeding. Similarly, if an infant is in diapers ($d_i = 1$), their rate of infectiousness to their mother becomes $D\tau_i$ where D incorporates the number of diaper changes per day and the increase in transmission due to a mother's exposure to diapers. Our model tracks which children belong to each mother so that these increased rates of transmission can be appropriately allocated to the correct individual. Note, we assume these increased risks of CMV transmission only apply to the mother of the child and not to another parent or caregiver.

Whether children are breastfeeding or in diapers is determined stochastically based on the age of the infant and survival curves for each behaviour. Data from Belgium on the age at which children are entirely toilet trained (55) was extracted from published figures using WebPlotDigitizer and used to fit a Gompertz survival function via nonlinear least squares. From this, we calculated the age-dependent transition probabilities for when children become toilet trained for each model time step. Similarly, other data from Belgium on the age at which children stop breastfeeding (56) was used to fit an exponential survival function to calculate similar transition probabilities for stopping breastfeeding. By stochastically drawing from these transition probabilities at every time step in model simulations, children in our model transition out of breastfeeding and/or out of diapers. As these transitions occur, the added risk of transmission between mothers and children is eliminated. Graphs showing data on toilet training and breastfeeding cessation are shown in Figure S. 1.

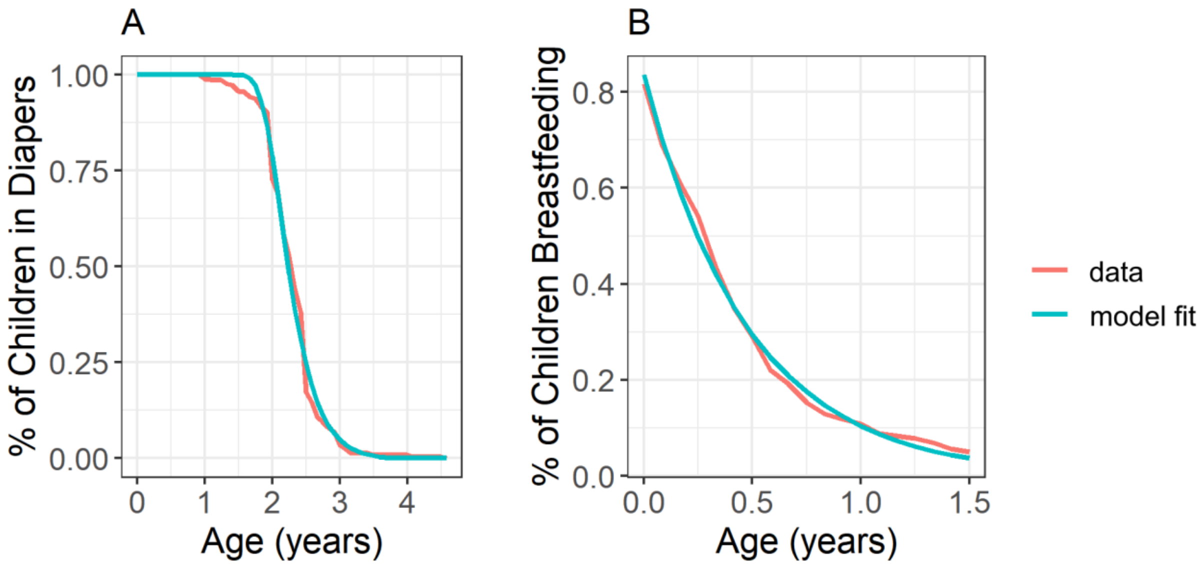


**Figure S. 1: Age-dependent change in the percentage of children in diapers and breastfeeding.** Data describing the percentage of children in diapers (**A**) comes from (55) while data describing the percentage of children breastfeeding (**B**) comes from (56). Fits to data were generated through by fitting a Gompertz survival curve and an exponential curve for **A** and **B**, respectively.

### 1.4 Immune protection

In addition to the infectiousness of a contact, the rate at which an individual becomes infected or reinfected is also based on the level of immunity of that individual, $\rho_i$, where $0 \leq \rho_i \leq 1$. Individuals in the susceptible class are assumed to have no protection, $\rho_i = 0$. Individuals with primary infection or reinfection are assumed to have waning protection, equal to $P_P(a_i, \Delta t_i)$ and $P_R(a_i, \Delta t_i)$, respectively. From this, we can say that the immune protection of individual i of infection status $s_i$ at time $\Delta t_i$ after the transition into their current infection stage is

$$\rho_i(a_i, \Delta t_i, s_i) = \begin{cases} 0 & , s_i = \text{uninfected} \\ P_P(a_i, \Delta t_i) & , s_i = \text{primary infected} \\ P_R(a_i, \Delta t_i) & , s_i = \text{reinfected.} \end{cases}$$

Immediately following primary infection or reinfection, we assume complete protection against reinfection, ($P_P(a_i, 0) = P_R(a_i, 0) = 1$). However, we assume this immune protection wanes at a rate $r_P$, until it reaches a constant, chronic level of protection, $P_C$.

We also assume that an individual with primary infection maintains complete protection ($\rho_i = 1$) against reinfection until the time $\Delta t = t^*$ when their infectiousness has waned to what it would be immediately following reinfection ($T_p(a_i, t^*) = T_R(a_i, 0)$, or $t^* = \ln(1/R)/r_T(a_i)$). This way, reinfection always ensures an increase in transmissibility. With this assumption, the level of protection following primary infection is

$$P_P(a_i, \Delta t_i) = \begin{cases} 1 & , \Delta t_i < t^* \\ e^{-r_P(\Delta t_i - t^*)} & , t^* \leq \Delta t_i < t^* + \dfrac{\ln(1/P_C)}{r_P} \\ P_C & , \Delta t_i \geq t^* + \dfrac{\ln(1/P_C)}{r_P} \end{cases}$$

and protection following reinfection is

$$P_R(a_i, \Delta t_i) = \begin{cases} e^{-r_P \Delta t_i} & , 0 \leq \Delta t_i < \dfrac{\ln(1/P_C)}{r_P} \\ P_C & , \Delta t_i \geq \dfrac{\ln(1/P_C)}{r_P}. \end{cases}$$

Graphs of $\mathbf{P_p(a_i, \Delta t_i)}$ and $\mathbf{P_R(a_i, \Delta t_i)}$ are shown in Figure 2 G of the main paper.

#### 1.5 Rate of acquisition

With the rate of contact, the per-contact infectiousness rate, and immune protection of each individual defined, the rate of infection acquisition for an individual i of age j and sex k through interactions with the population becomes

$$\beta_{P,i,j,k} = (1 - \rho_i(a_i, \Delta t_i, s_i)) \cdot$$

$$\sum_{age,\ell} \sum_{sex,m} \frac{\sum_{\forall \text{ individual,n of age,}\ell \text{ and sex,m}} \tau_n(a_n, \Delta t_n, s_n)}{N_{m,\ell}} \zeta_{j,k,\ell,m}$$

where $N_{\ell,m}$ is the number of individuals in the population of age $\ell$ and of sex m, and $\zeta_{j,k,\ell,m}$ is the per-unit time contact rate that an individual of age group j and sex k has with other individuals of age $\ell$ and sex m, according to the contact matrix. In this way, we take the average per-contact transmission rate across each age and sex group.

An individual's rate of infection acquisition through interactions with their set of children $V_i$ is

$$\beta_{C,i,V_i} = \begin{cases} 0 & , \sigma_i = 0 \\ (1 - \rho_i(a_i, \Delta t_i, s_i)) \displaystyle\sum_{\forall\, v \epsilon V_i} d_v D \tau_v(a_v, \Delta t_v, s_v) & , \sigma_i = 1 \end{cases}$$

where $\sigma_i \epsilon \{0,1\}$ and $\sigma_i = 0$ indicates the individual i not a mother and $\sigma_i = 1$ indicates individual i is a mother.

The rate of infection acquisition for individual i with mother x through breastfeeding is

$$\beta_{B,i,x} = \begin{cases} 0 & , b_i = 0 \\ (1 - \rho_i(a_i, \Delta t_i, s_i)) B \tau_x(a_x, \Delta t_x, s_x) & , b_i = 1. \end{cases}$$

Together, the overall per unit time rate of transmission for individual i is

$$\beta_i = \beta_{P,i,j,k} + \beta_{C,i,V_i} + \beta_{B,i,x}.$$

### 1.6 Aging and births

We track individuals throughout their lives from birth, deterministically aging them at each model time step until they reach an age of $a_{max} = 50$ years where they exit the model. We chose a maximum age of 50 years as past this point individuals are assumed to have decreased contact with children and women are assumed to have exited their child-bearing years. We assume women in the population give birth at an age-dependent rate based on values in Statistics Canada's 2016 Demographics Report (57). This report breaks women into five-year age groups and reports the birth rate for each group. With these assumed birth rates, women have an average of 1.54 children over the course of their lifetime. This birth rate is below replacement. Because we do not include the impact of death before the age of 50, immigration, or emigration in our model, birth rate alone determines the change in population size over time. To ensure a steady population size, we scale the birth rates to ensure that, on average, women have 2 children over the course of their lifetime. Both the original birth rates reported in (57) and the adjusted values used in our model can be found in Table S. 1.

| Age Group | Birth rate per woman per year | Scaled birth rate per woman per year |
|---|---|---|
| 15-19 | 0.0084 | 0.0109 |
| 20-24 | 0.0376 | 0.0488 |
| 25-29 | 0.0876 | 0.1138 |
| 30-34 | 0.1076 | 0.1397 |
| 35-39 | 0.0556 | 0.0722 |
| 40-44 | 0.0115 | 0.0149 |

**Table S. 1: Birth rates of women according to Statistics Canada.** These values were scaled to ensure that women have an average of 2 children over the course of their lifetime resulting in a steady population size.

When mothers give birth, the infant's CMV infection status depends on the CMV infection status of the mother over the last nine months of pregnancy. The probability of congenital CMV transmission is directly linked to their maximum infectiousness over the course of pregnancy. Mothers who acquire a primary infection during pregnancy are at the highest risk of passing on congenital CMV to their child with a probability of $\phi = 0.334$ (33). As the infectiousness of the mother declines, so does her risk of transmitting CMV to her fetus. Thus, the probability of mother i passing cCMV to her child is the maximum value of

$$\phi \cdot \frac{1}{T(a_i)} \cdot \tau_i(a_i, \Delta t_i, s_i)$$

over the nine months of her pregnancy.

Both the selection of a woman to give birth and the selection of the CMV infection status of the child at birth are randomly drawn from an Euler-multinomial distribution based on the above probabilities, determined for each time step in the model.

### 2. Model fitting

Parameters of our model were fit using ABC, following the Lenormand Algorithm (38). Relating to rates of transmission, we use this algorithm to determine the infectiousness of children and adults immediately following primary infection (parameters $C$ and $A$, respectively), the infectiousness of individuals in the chronic stage of infection (parameter $T_C$) and the factor by which infectiousness is reduced during reinfection compared to primary infection (parameter $R$). Pertaining to immunity, we assume immune protection takes on a value between 0 and 1 and can be thought of as the fraction reduction in an individual's likelihood of infection due to immunity. Susceptible individuals have an immune protection of 0, while newly infected individuals have an immune protection of 1. We fit parameter $r_P$, the rate at which immunity wanes following a new infection, and $P_C$, the level of immune protection possessed by individuals in the chronic stage of infection. Finally, to capture the transmission rate due to mothers' interactions with their infants' diapers and infants' interactions with infectious breastmilk, we fit parameters $D$ and $B$, respectively.

Values for the parameters being fit are randomly selected and used to simulate the model. From this simulation, we calculate summary statistics and compare these values to equivalent statistics in available data. How well the summary statistics of the simulation match with the summary statistics in of the data informs the next selection of parameter values. This continues until the selected values for each parameter of the model converge.

We used the percentage of children born with congenital CMV, and the percentage of individuals with CMV at every age from 1-49 as our summary statistics. The target percentage of children born with congenital CMV was to set equal 0.6% from data out of the United States of America (US) (40). The proportion of individuals with CMV at ages 1-49 came from data from the NHANES, a series of surveys conducted by the National Center for Health Statistics for the Center of Disease Control in the US. As part of this study, participants provided serum that was tested for the presence of anti-CMV IgG antibodies via ELISA. We pooled data collected during the survey years of 1988-1994, 1999-2000, 2001-2002, 2003-2004, and 2011-2012. A full analysis of these data can be found in (58,59). All data is publicly available and can be found at https://wwwn.cdc.gov/nchs/nhanes/Default.aspx.

Simulations were run for 70 years, with the last 20 years used to calculate these summary statistics. These values were compared to the known occurrence of cCMV and the age-specific prevalence of CMV, broken into years, as described in the US NHANES data set by calculating the average of the percent error for each statistic.

### 3. Defining the effective reproduction number

When simulating different vaccine strategies, we calculated the change in the effective reproductive number ($R_{eff}$), in individuals in the years following vaccine implementation. To calculate this value, we set $R_{eff}$ to be

$$R_{eff}(t) = \frac{\mu(t)[\alpha_{max} - \alpha(t)]}{\eta(t)}.$$

Here, at time $t$, $\mu(t)$ is the expected number of new infections per year, $\eta(t)$ is the number of infected individuals in the population, and $\alpha(t)$ is the mean age of CMV infection acquisition. Because CMV is a lifelong infection and is maintained until individuals age out of the model (age of $\alpha_{max} = 50$), the total length of time during which an individual could cause new infection is $\alpha_{max} - \alpha(t)$.

### 4. Impact of a CMV vaccine inducing sterilizing immunity

We simulated the impact of an idealized vaccine that provides complete, life-long sterilizing immunity when given to different age groups and different fractions of the population. As the likelihood of cCMV is directly determined by the infection status of the mother, we examined how the distribution of pregnant women's infection statuses changed following vaccine implementation (Figure S. 2). Changes in $R_{eff}$ in the years following vaccine implementation are shown in and Figure S. 3.

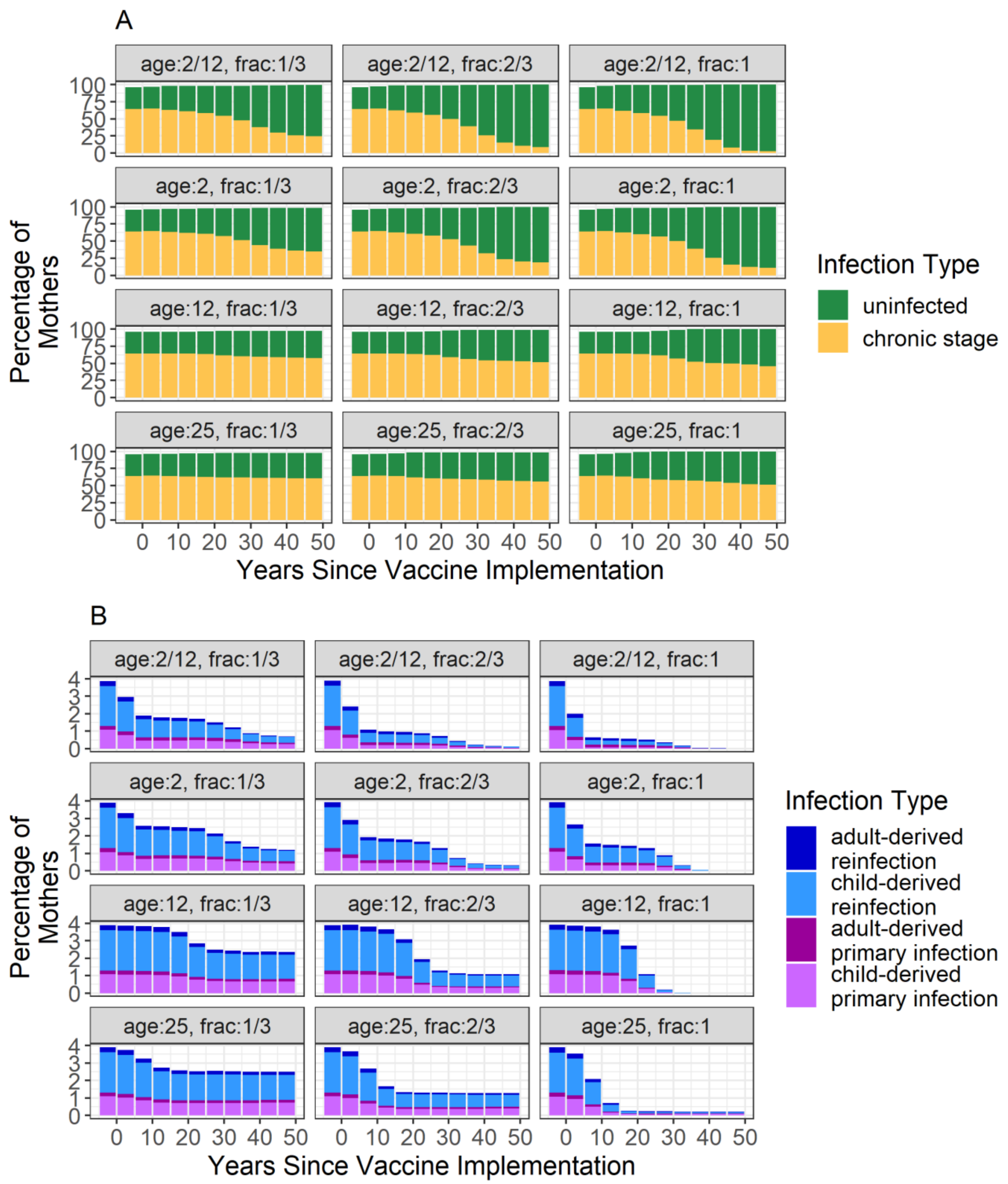


**Figure S. 2: CMV infection-stage and prevalence in pregnant women following implementation of an idealized vaccine that provides sterilizing immunity.** Figures are stratified by the fraction of the population vaccinated, and the age in years when the vaccine is given. The change in the fraction of women who are uninfected or whose infectiousness is at the chronic level throughout pregnancy is shown in **panel A.** The change in the fraction of women who acquire a primary infection or reinfection during pregnancy is shown in **panel B.**

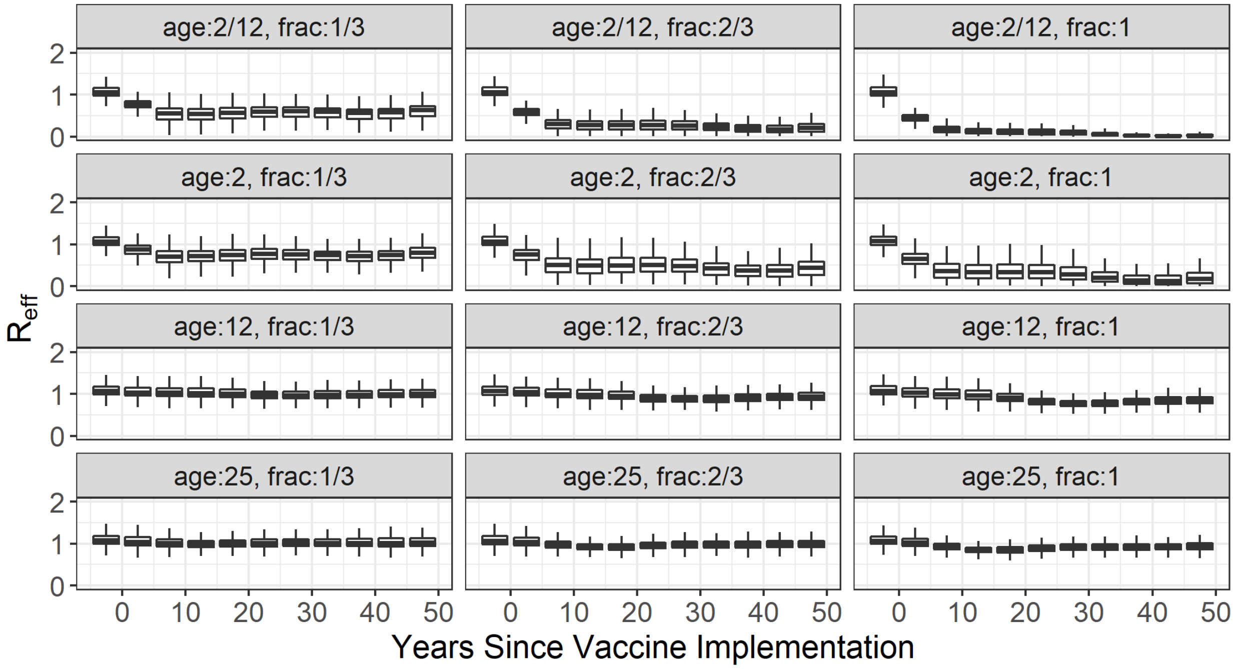


**Figure S. 3: Decline in the $R_{eff}$ following implementation of a vaccine providing sterilizing immunity.** Younger ages of vaccination lead to faster and more dramatic declines in $\mathbf{R_{eff}}$. The figure is stratified by the fraction of the population vaccinated and the age in years at which vaccines are given.

### 5. Impact of a vaccine inducing natural protection

When simulating the impact of a vaccine inducing the equivalent of natural protection, we again examined how such a vaccine would change the infection statuses of pregnant women and the $R_{eff}$. Graphs of these values are shown in Figure S. 4 and Figure S. 5, respectively.

The impact of such a vaccine on the infection statuses and infectiousness of children is shown in Figure S. 6. Following vaccination, vaccinated children can still have breakthrough infections (Figure S. 6 A); however, vaccination allows the infectiousness of infected children, and thus how likely they are to transmit infection to others, to decline (Figure S. 6 B).

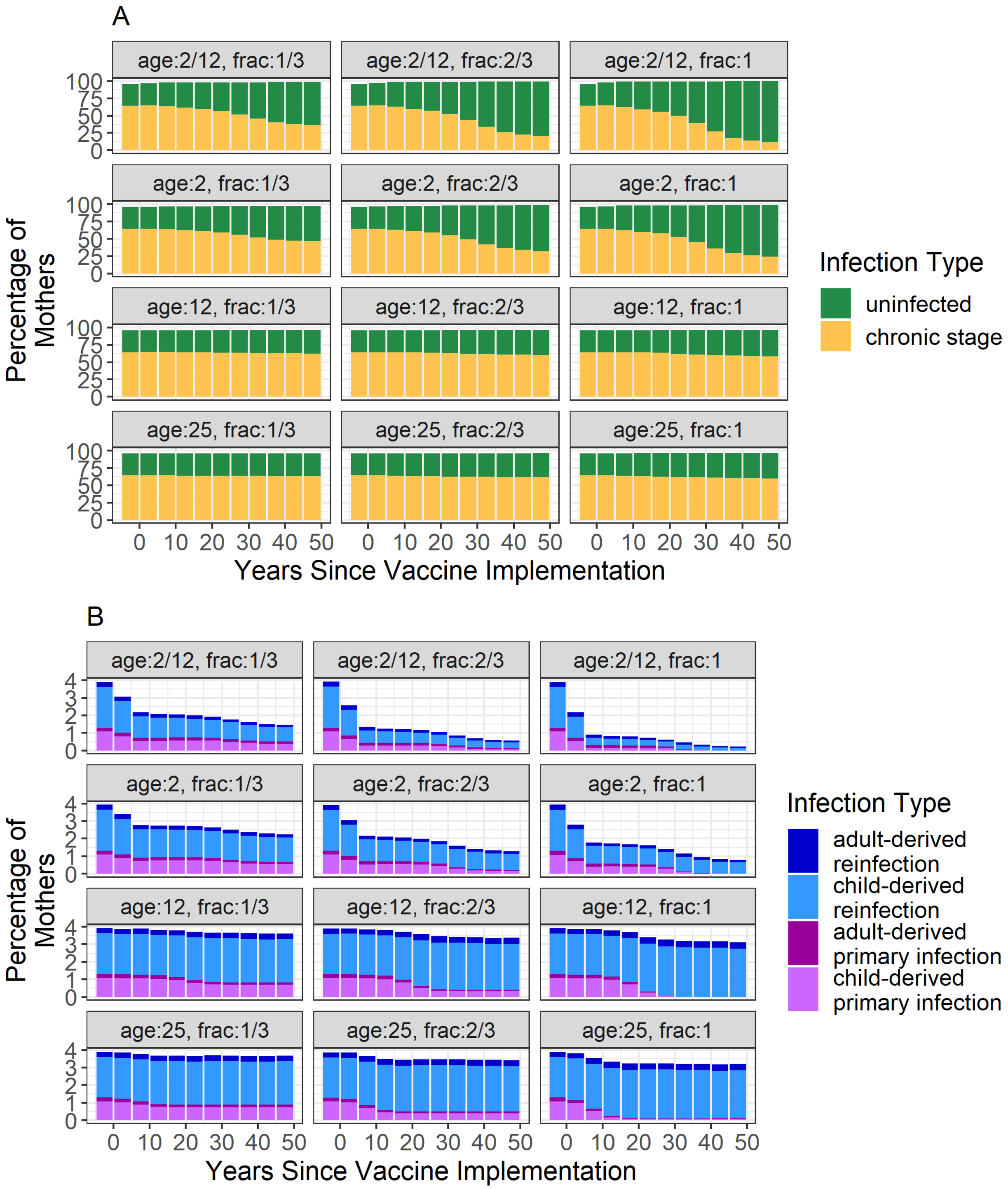


**Figure S. 4: CMV infection-stage and prevalence in pregnant women following implementation of a vaccine that provides natural immunity.** Bars show the mean prevalence, as predicted by model simulations, and are stratified by the fraction of the population vaccinated, and the age in years vaccines are given. The change in the numbers of women who are uninfected or whose infectiousness is in the chronic stage throughout pregnancy is shown in **panel A**. The change in the numbers of women who a primary infection or reinfection during pregnancy is shown in **panel B.** Note that vaccinated women acquiring their first infection during pregnancy are classified here as having a reinfection.

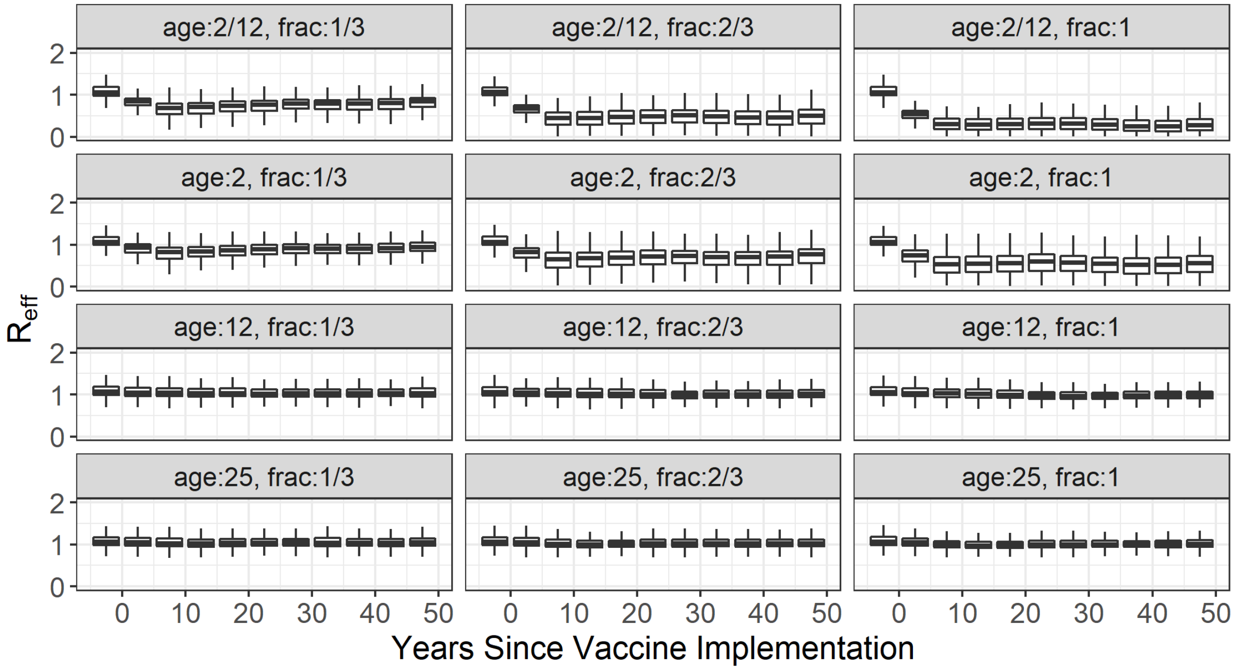


**Figure S. 5: Decline in the $R_{eff}$ following implementation of a vaccine that provides natural immunity.** Younger ages at time of vaccination lead to faster and more dramatic declines in $\mathbf{R_{eff}}$. The figure is stratified by the fraction of the population vaccinated and the age in years at which vaccines are given.

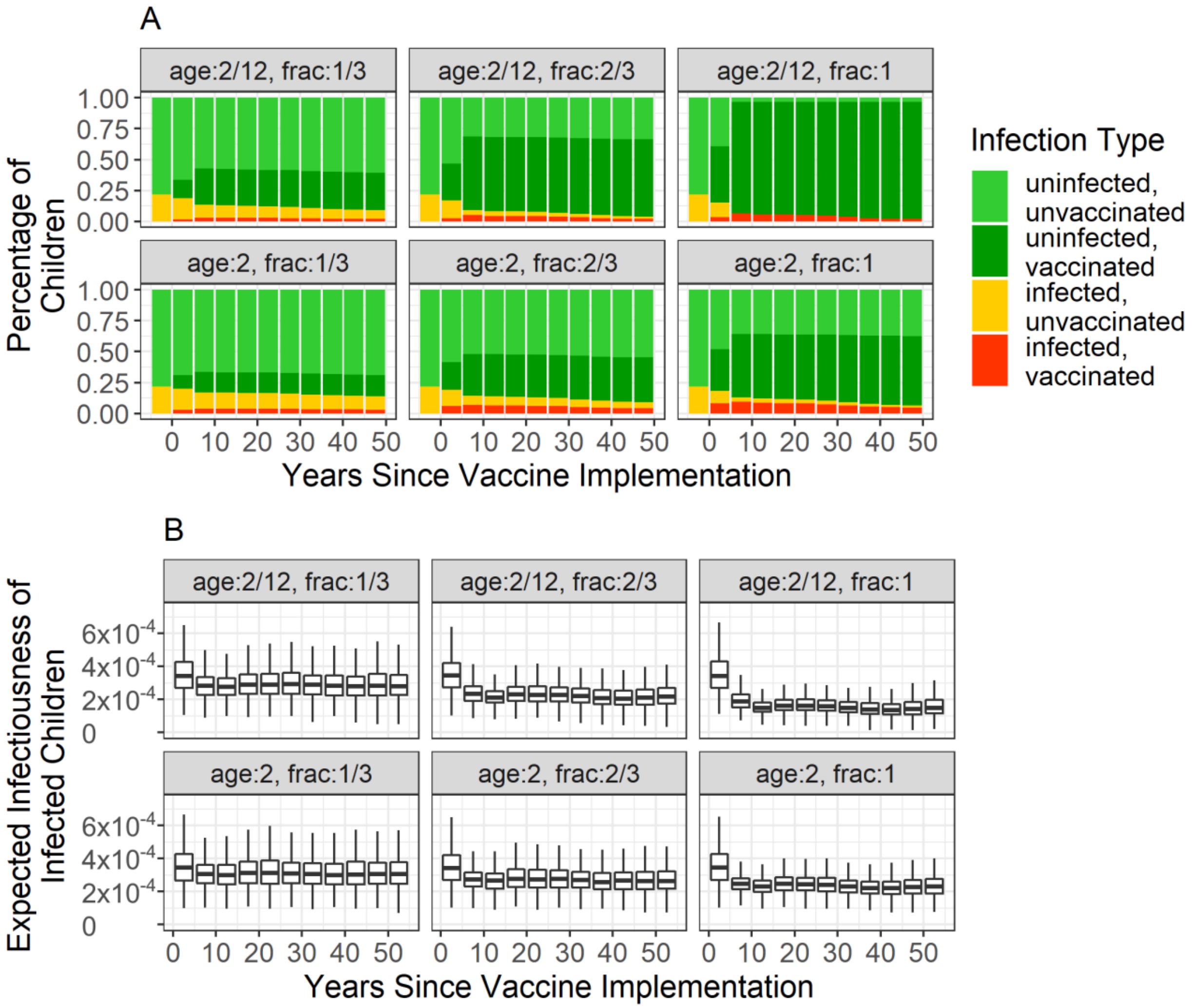


**Figure S. 6: Impact of a vaccine mimicking natural immunity on children's infection status and infectiousness. Panel A** shows the percentage of children (≤ 5 years old) infected with CMV, with and without vaccination, in the years following vaccine implementation. In **panel B**, expected infectiousness of infected children (≤ 5 years old) in the years following vaccine implementation is shown. Graphs are stratified by the age of vaccine deliverance in years and the fraction of vaccine uptake in the population. Because vaccination does not occur until either 2-months-old or 2-years-old, even with a vaccine uptake fraction of 1, not all children at any given time (which includes those under the age of vaccination) will be vaccinated.

### 6. Effects of booster shots when using a vaccine that invokes natural immunity

When analyzing the impact of a vaccine that invokes natural immunity, we simulated delivering booster shots at various time intervals following initial vaccination. Results are shown in Figure S. 7. None of the booster shot scenarios significantly reduced cCMV cases, when compared to a one-shot scenario.

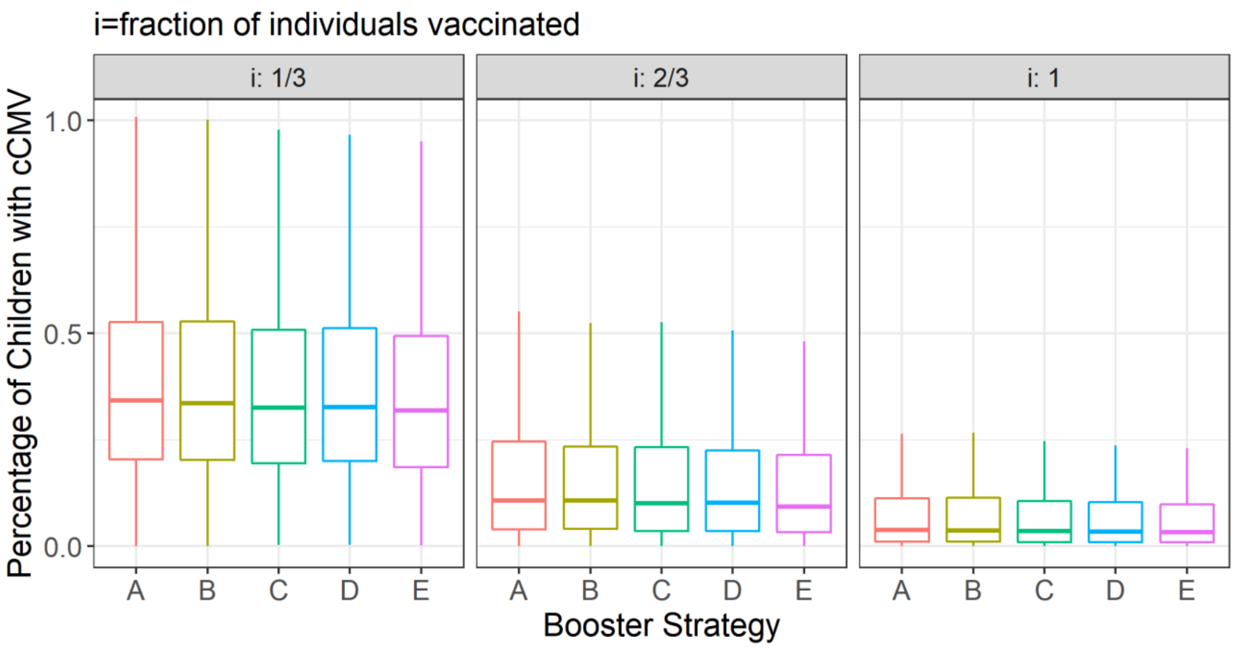


**Figure S. 7: Decrease in the prevalence of cCMV when using booster shots.** All graphs show the predicted percentage of children born with cCMV 50 years after implementation of a vaccine that mimics natural immunity given to 2-month-olds. Booster strategy A represents what occurs when only a single vaccine dose is administered. Booster strategy B represents the addition of a booster at 2 years of age. Booster strategy C represents what occurs when the booster is given at 12 years. Booster strategy D represents what occurs when a booster is given at X years, at which time vaccine-induced immunity has waned to half its initial value (after 275 days, on average). Booster strategy E represents what occurs when 2 boosters are given, at X and Y years, at which times vaccine-induced immunity wanes to half its initial value. In all scenarios in which boosters are included (B-E), no substantive decreases in cCMV cases are apparent.

4. Rosenthal LS, Fowler KB, Boppana SB, Britt WJ, Pass RF, Schmid SD, et al. Cytomegalovirus shedding and delayed sensorineural hearing loss: results from longitudinal

follow-up of children with congenital infection. *Pediatric Infectious Disease Journal*. 2009;28(6): 515–520. https://doi.org/10.1097/INF.0b013e318198c724.
